# Associations Between Female Sex Hormones and Skeletal Muscle Ageing: The Baltimore Longitudinal Study of Aging

**DOI:** 10.1101/2024.10.06.24314971

**Authors:** Annabel J. Critchlow, Sarah E. Alexander, Danielle Hiam, Luigi Ferrucci, David Scott, Séverine Lamon

**Affiliations:** School of Exercise and Nutrition Sciences, Institute for Physical Activity and Nutrition (IPAN), Deakin University, Geelong, Australia; Cardiometabolic Health and Exercise Physiology, Baker Heart and Diabetes Institute, Melbourne, Australia; School of Clinical Sciences at Monash Health, Monash University, Clayton, Australia; National Institute on Aging, National Institutes of Health, Baltimore, Maryland, United States

**Author notes:** **Corresponding author:** A/Prof. Séverine Lamon, School of Exercise and Nutrition Sciences, Faculty of Health, Deakin University, 221 Burwood Hwy, Burwood 3125., Australia, Ph. **Author contact details:** Annabel Critchlow, Dr Sarah Alexander, Dr Danielle Hiam, Dr Luigi Ferruci, A/Prof David Scott.

**Keywords:** oestrogen, ovarian hormones, skeletal muscle, ageing

## Abstract

**Background:** To date, most research investigating the influence of circulating sex hormones on ageing female skeletal muscle has been cross-sectional and focused only on dichotomised young and old, or pre-versus post-menopausal groups. This excludes an important transitional period from high to low circulating oestrogen. Using secondary data from the Baltimore Longitudinal Study of Aging, this study aimed to investigate cross-sectional and longitudinal associations between circulating sex hormones and skeletal muscle mass and function across a continuum of ages.

**Methods:** Multiple and binomial linear regression was used to map cross-sectional (n=319) and longitudinal (n=83) associations between circulating sex hormones (oestradiol (E2), free oestradiol index (FEI), free (TT) and bioavailable testosterone (BioT), testosterone/oestradiol ratio (TT/E2)) and skeletal muscle mass and function in healthy females. Cross-sectional models analysed females across an ageing continuum (24-89 years) and longitudinal associations were tested across 4-6 years of ageing in females over 50 years old. Models were adjusted for age, height, physical activity, comorbidities, and ethnicity.

**Results:** Cross-sectionally, serum E2 and FEI were positively associated with relative appendicular lean mass (ALM; β=0.28 and 0.20, respectively, *p*<0.05) and thigh muscle percentage (β=0.19 and 0.15, respectively, *p*<0.05). E2 and FEI were negatively associated with total body fat percentage (β=-0.30 and -0.21, respectively, *p*<0.05). BioT was positively associated with absolute ALM (β=0.13, *p*<0.05) and total body fat percentage (β=0.18, *p*<0.05). TT was negatively associated with total body fat percentage (β=-0.14, *p*<0.05). The TT/E2 ratio was negatively associated with thigh muscle CSA (β=-0.08, *p*<0.05) and hamstring strength (β=-0.12, *p*<0.05). Across 4-6 years, a decrease in E2 and FEI were associated with a decrease in ALM (β=0.29 and 0.42, respectively, *p*<0.05), and decrease in FEI was associated with a decrease in handgrip strength (β=0.17, *p*<0.05). A decrease in TT and BioT were associated with an increase in relative ALM (β=-0.25 for both, *p*<0.05) and relative quadriceps strength (β=-0.12 and -0.27, respectively, *p*<0.05)

**Conclusion:** This study demonstrates novel associations between sex hormone levels and skeletal muscle in females across a wide continuum of ages. We also demonstrate that longitudinal fluctuations in circulating sex hormones must be considered to gain a comprehensive understanding of female muscle ageing.

## Introduction

The World Health Organization [1] predicts the global population above 60 years of age will double from 2019 to 2050. However, improvements to increase the lifespan have not been matched by advancements in the health span; that is, the length of time a person remains healthy, not just alive. While females live longer than males [2], those additional years are spent in significantly poorer health, highlighting a large disparity between the health span and lifespan in females compared to males [3].

Ageing is associated with a progressive loss of muscle mass and function which presents an increasingly substantial public health concern. Age-related muscle wasting is significantly associated with many adverse outcomes, including reduced independence and quality of life, and increased chronic disease, hospitalisation, and mortality [4]. While the prevalence of age-related muscle loss is seemingly similar between the sexes [5], evidence suggests that the patterns, and therefore the underlying molecular mechanisms of muscle ageing may be sex-specific. For example, females undergo a rapid decline in knee extensor strength ∼20 years earlier than males [6]. In addition, while both sexes share the same direction of changes to the transcriptome with ageing, the magnitudes of change to differentially expressed genes differs between the sexes from young and old individuals [7].

Testosterone and oestrogen are the predominant sex hormones in males and females, respectively. While they are produced by both sexes, their profiles vary drastically across the lifespan. Females undergo a dramatic decline in endogenous oestrogen production starting ∼40 years of age [8], meaning they can spend over a 3^rd^ of their life in an oestrogen-deficient state. A steep reduction in total and bioavailable (i.e. not bound to sex hormone-binding globulin (SHBG)) testosterone occurs in the early reproductive years, which stabilises around menopause [9]. In contrast, male testosterone production remains largely consistent across adult life with a gradual decline in later decades [9]. Oestrogen and testosterone elicit effects in multiple tissues, including skeletal muscle, by binding to the oestrogen (ER) and androgen receptor (AR), respectively [10]. In females, bioavailable testosterone is associated the maintenance of muscle mass [11], and oestrogen may regulate the maintenance of muscle mass, strength, and mitochondrial function [12].

A recent systematic review by our group found a consistent reduction in muscle mass and handgrip strength across the menopausal transition, however many studies show no association between oestradiol concentrations (E2, the major endogenous form of oestrogen) and muscle mass or function in pre- or postmenopausal females [13]. The cross-sectional nature and dichotomisation of age groups in this body of evidence is a limitation because it prevents conclusions to be drawn about the effects of changes in sex hormone concentrations on muscle health over time. Only one study has explored this association in a longitudinal manner, reporting no relationship between baseline circulating E2 or testosterone and the loss of appendicular lean mass or knee extensor strength over 3 years in postmenopausal females [14]. However, it remains largely unknown how age-related fluctuations in sex hormones impact the regulation of skeletal muscle across a broader age range, as associations between the changes in sex hormones and skeletal muscle health across ageing and the menopausal transition are still to be mapped.

This study used data from the Baltimore Longitudinal Study of Aging (BLSA, [15]) to address two research questions. First, we aimed to determine the cross-sectional associations between circulating female sex hormone concentrations and skeletal muscle mass and function across a continuum of ages and hormonal status (24 to 89 years). Secondly, we aimed to investigate the longitudinal associations between circulating sex hormone concentrations and the changes in muscle mass and function that occurred over a 4–6-year period in females aged 50 to 87 years.

## Methods

### Study Population

This is a secondary analysis of data from the BLSA [15], an ongoing prospective study that aims to extensively map the process of healthy ageing [15]. Testing is carried out at the National Institute of Ageing Clinical Research Unit in Baltimore, Maryland. Healthy participants between 20-96 years without any chronic disease or cognitive impairment were recruited. Data collected between 2003 and 2023 were utilised in this analysis. Participants under 60 years old were tested every 4 years, 60–79-year-olds were tested every 2 years, and those above 80 years were tested every year.

#### Inclusion Criteria

Inclusion criteria for the present study included female BLSA participants with no missing variables of interest (serum sex hormones, DXA body composition, handgrip, hamstring, and quadriceps strength, and confounding variables). Data from females at their first visit was used for the cross-sectional analysis if there were no missing variables (n=319). From these participants, females >50 years were included in the longitudinal analysis (n=83) if they had a follow up visit 4-6 years later with all the required sex hormone and outcome variables. If they had multiple visits within this time frame, data from the latest was used. As the BLSA recruit only healthy individuals at baseline, there were no additional exclusion criteria specific to this study.

### Measures

#### Serum Hormone Analysis

Fasted (12 h) blood samples were taken from participants in the morning of each visit and immediately centrifuged at 4°C. Serum was aliquoted and stored at -80°C. Fasted serum hormone concentrations of E2 (ng/dL) and total testosterone (TT, ng/dL) were determined by high-performance liquid chromatography-tandem mass spectrometry (LC/MS; Esoterix part of LabCorp, Calabasas Hills, CA). SHBG concentrations (nmol/L) were analysed via immunoradiometric assay (Esoterix part of LabCorp). To determine bioavailable testosterone (BioT, ng/dL), ammonium sulphate was used to separate SHBG-bound hormones from albumin-bound and free hormones (Esoterix part of LabCorp). E2, TT, and BioT concentrations were converted to SI units (pmol/L, nmol/L and nmol/L, respectively). The free oestrogen index (FEI) was calculated by dividing E2 (pmol/L) by SHBG (nmol/L) and multiplying by 100. The testosterone to oestrogen (TT/E2) ratio was calculated by dividing TT (nmol/L) by E2 (pmol/L).

#### Body Composition

Body composition measures including appendicular lean mass (ALM, kg), leg lean mass (LLM, kg), and total body fat mass (TBFM, kg) were assessed via whole-body DXA (Prodigy Scanner, GE, Madison, WI) scans with software version 10.51.006. The scanner was calibrated daily. Reliability has been assessed in 12 older (>65 years) males, showing <1% difference in fat mass (kg) between 2 scans, 6 weeks apart [16]. Relative ALM (kg.kg^-1^) was calculated by dividing appendicular lean mass (kg) by total body fat mass (kg). Thigh muscle cross-sectional area (CSA, cm^2^), subcutaneous fat area (cm^2^), intramuscular fat area (cm^2^), and muscle density (HU) were assessed using computerised tomography (CT) cross-sectional images of the mid-thigh (Somatom Sensation 10, Siemens, Malvern, PA). Thigh muscle percentage was calculated by dividing total thigh CSA (cm^2^) by muscle CSA (cm^2^). CT images were analysed and quantified with Geanie 2.1 software (BonAlyse Oy, Jyvaskla, Finland).

#### Muscle Function

Handgrip strength (kg) was assessed using a Jamar Hydraulic hand dynamometer (Patterson Medical, Warrenville, IL). Participants sat with their arm extended to 180° or shoulder height and were instructed to squeeze the dynamometer as hard as possible. Maximum force was measured in three trials on each hand, and the best of 6 were used in the analysis. The average of the left and right hand was used in the analysis. Lower limb strength was measured using the Kin-Com isokinetic dynamometer (Kin-Com model 125E version 3.2, Chattanooga Group, Chattanooga, TN) from 2003 to 2011, and the Biodex Multi-Joint System-Pro dynamometer (Biodex Medical System, Advantage Software V4.X, Inc., Shirley, NY) from 2011 to 2023. Quadriceps strength (isokinetic knee extension; Nm) and hamstring strength (isokinetic knee flexion; Nm) was measured at an angular velocity of 30°/s and 180°/s. Participants were asked to extend/flex their knee as hard as possible through a range of motion set between 100-160°. They completed two practice trials, followed by three test trials. The highest peak torque value was included in the analysis. Peak torque normalised to leg lean mass (Nm/kg) and thigh muscle CSA (Nm/cm^2^), were calculated by dividing hamstring and quadriceps peak torque (Nm) by leg lean mass (kg) and thigh muscle CSA (cm^2^), respectively.

#### Physical Function

Gait speed was assessed by instructing participants to walk 400 m as fast as possible after a warm-up of 2.5 minutes at a usual gait speed. Chair stand ability was measured by asking participants to fold their arms and stand up from a chair 10 times. The pace of the first 5 chair stands (stands/s) was recorded. Participants completed three tests (6 m walk test, chair stand test, and standing balance) to comprise the Short Physical Performance Battery (SPPB), a validated assessment of mobility. Performance in each of the 3 tests was scored from 0 (worst) to 4 (best) and summed to give a total out of 12, as previously described [17]. Finally, participants were categorised as a faller if they had at least one self-reported fall in the previous 12 months.

#### Potential confounding variables

Demographic information was collected from participants via an interview, including age, ethnicity, comorbidities, and total physical activity in the previous two weeks. The age adjusted Charlson comorbidity index (ACCI) was determined as previously described [18]. Briefly, a score is assigned to each comorbid condition, with additional points for increasing age groups.

### Statistical Analysis

Statistical analysis was completed using Stata software version 17.0 (StataCorp, College Station, TX). Data are presented as mean ± SD and statistical significance was accepted as p<0.05. Participants with values that were deemed physiologically implausible (>10 SDs) were excluded. One participant was excluded from the cross-sectional analysis because handgrip strength was >300kg. Forty and 21 participants were excluded from the cross-sectional and longitudinal analysis, respectively, because their hormone levels were below zero. Paired t-tests were performed to determine the differences between variables at baseline and follow-up.

#### Multiple regression

Multiple linear regression was performed to determine the cross-sectional relationships between circulating sex hormones, body composition (from DXA), and muscle function. Further outcome variables were assessed in subsets of participants where measured, including thigh CT measures and physical function. Additional models were performed to determine cross-sectional associations between self-reported menopausal and hormonal supplementation status and all muscle outcomes. In the longitudinal analysis, multiple regression models were performed to determine the relationship between the change in sex hormone levels and the change in outcome measures across 4-6 years in females over 50 years. Further analysis was completed to measure the longitudinal associations between baseline menopausal status and HRT use and the change in muscle outcome measures.

Each model was adjusted for age, ethnicity, height, ACCI, and physical activity. Where significant and opposing associations were detected for serum oestradiol (E2 or FEI) and testosterone (TT or BioT), both hormones were added to the model to determine if they remained significantly and independently associated with the outcome variable.

Multicollinearity was assessed by variation inflation factors with a threshold of 5. No variables displayed collinearity. Normality of residuals was confirmed by plotting a histogram of studentised residuals. Homoscedasticity was assessed by plotting fitted versus predicted residuals. Some heteroscedasticity was observed, so robust standard errors were used. To determine the magnitude of the relationships, the z-score was calculated for all independent (E2, FEI, TT, BioT, SHBG, and TT/E2 ratio) and outcome measures (body composition, muscle function, and physical function), where zero equals the mean, and 1 equals the standard deviation. Where the standardised coefficient was <0.2, the effect size of the relationship was considered ‘small’, 0.2 to 0.5 denotes a ‘medium’ effect size, and >0.5 represents a ‘large’ effect size.

#### Binomial linear regression

In females over 50 years of age, binomial linear regression models were used to determine the cross-sectional relationship between sex hormone levels and dichotomous physical function outcome measures (mobility disability and fallers).

## Results

### Participant characteristics

Characteristics of the participants included in the cross-sectional and longitudinal analysis are shown in Tables 1 and 2, respectively. Self-reported menopausal status and hormonal supplementation is shown in Supplementary Table S1. In the longitudinal analysis, there was a 50.8% reduction (*p*<0.05) from baseline (mean age: 64.41 ± 10.99 years) to follow up (4.86 ± 0.88 years later) in circulating E2, but not in other hormones. In body composition measures, there was a 4.6% reduction in thigh muscle CSA (*p*<0.05), and a 3.9% reduction in thigh muscle density (*p*<0.05). Regarding muscle function, there was a reduction in absolute and relative hamstring strength at 180 °/s (absolute: -12%, relative to LLM: -11.2%, relative to muscle CSA: -8.4%, *p*<0.05), and absolute and relative quadriceps strength at 180°/s (absolute: -8.0%, relative to LLM: -7.5%, *p*<0.05) and 30°/s (absolute: -8.2%, relative to LLM: -8.1%, relative to CSA: -4.5%, *p*<0.05). There was a 9% increase in 400 m walk time (*p*<0.05).

**Table 1.** Participant characteristics from cross-sectional analysis.

| Participant Characteristics | n | Mean | SD | Range |
| --- | --- | --- | --- | --- |
| **General** |  |  |  |  |
| Age (years) | 319 | 60.20 | 15.95 | 24.00 - 89.00 |
| **Sex hormone levels** |  |  |  |  |
| E2 (pmol/L) | 319 | 122.78 | 234.64 | 5.14 - 1872.21 |
| FEI | 319 | 0.16 | 0.26 | 0.00 - 1.69 |
| TT (nmol/L) | 319 | 0.83 | 0.53 | 0.14 - 4.93 |
| BioT (nmol/L) | 319 | 0.13 | 0.10 | 0.02 - 0.80 |
| SHBG (nmol/L) | 319 | 81.65 | 45.49 | 13.87 - 379.00 |
| TT/E2 ratio | 319 | 33.43 | 30.55 | 0.45 - 158.33 |
| **Body composition** |  |  |  |  |
| BMI (kg/m<sup>2</sup>) | 319 | 26.51 | 5.02 | 17.83 - 45.74 |
| ALM (kg) | 319 | 17.49 | 2.78 | 10.93 - 25.76 |
| Total body fat (%) | 319 | 38.08 | 7.72 | 15.07 - 58.19 |
| Relative appendicular lean mass (kg/kg total fat) | 319 | 0.73 | 0.32 | 0.29 - 2.66 |
| thigh muscle CSA (cm<sup>2</sup>) | 239 | 92.53 | 22.43 | 51.99 - 256.99 |
| Thigh subcutaneous fat area (cm<sup>2</sup>) | 239 | 105.49 | 44.65 | 37.16 - 291.15 |
| Thigh intramuscular fat area (cm<sup>2</sup>) | 239 | 10.31 | 4.18 | 1.17 - 29.45 |
| Thigh muscle percentage (%) | 239 | 41.60 | 8.10 | 22.95 - 63.96 |
| Thigh muscle density (HU) | 239 | 51.02 | 3.66 | 38.65 - 60.85 |
| **Muscle function** |  |  |  |  |
| Hand grip strength (kg) | 319 | 25.56 | 6.83 | 10.00 - 53.00 |
| Hamstring peak torque (Nm) |  |  |  |  |
| 180d/s | 319 | 49.12 | 14.69 | 20.16 - 102.70 |
| 30d/s | 319 | 46.89 | 15.11 | 11.30 - 106.80 |
| Quadriceps peak torque (Nm) |  |  |  |  |
| 180d/s | 319 | 63.07 | 21.73 | 23.70 - 120.40 |
| 30d/s | 319 | 101.87 | 31.89 | 33.31 - 211.30 |
| Hamstring relative peak torque to LLM (Nm/kg) |  |  |  |  |
| 180d/s | 319 | 3.66 | 0.95 | 1.82 - 8.17 |
| 30d/s | 319 | 3.49 | 0.98 | 0.81 - 8.02 |
| Quadriceps relative peak torque to LLM (Nm/kg) |  |  |  |  |
| 180d/s | 319 | 4.68 | 1.40 | 1.56 - 9.66 |
| 30d/s | 319 | 7.56 | 1.97 | 2.19 - 13.21 |
| Hamstring relative peak torque to muscle CSA (Nm/cm<sup>2</sup>) |  |  |  |  |
| 180d/s | 239 | 0.53 | 0.12 | 0.22 - 0.90 |
| 30d/s | 239 | 0.51 | 0.14 | 0.13 - 0.93 |
| Quadriceps relative peak torque to muscle CSA (Nm/cm<sup>2</sup>) |  |  |  |  |
| 180d/s | 239 | 0.69 | 0.18 | 0.21 - 1.29 |
| 30d/s | 239 | 1.13 | 0.28 | 0.46 - 1.79 |
| **Physical function** |  |  |  |  |
| 6-metre walk time (s) | 203 | 5.17 | 0.91 | 3.31-7.92 |
| 400-metre walk time (s) | 163 | 277.40 | 49.70 | 199.88 - 531.61 |
| Time taken to complete 5 chair stands (s) | 203 | 9.50 | 2.70 | 3.57 - 18.52 |
| Mobility disability (%(n)) | 238 |  |  |  |
| Yes (SPPB ≤ 9) |  | 1.3 (3) | - | - |
| No (SPPB > 9) |  | 98.7 (235) | - | - |
| Falls in last 12 months (% (n)) | 238 |  |  |  |
| No falls |  | 80.7 (192) | - | - |
| Falls |  | 19.3 (46) | - | - |

Physical function measures were only assessed in females ≥ 50 years old. ALM: appendicular lean mass; BioT: bioavailable testosterone; BMI: body mass index; CSA: cross-sectional area; E2: oestradiol; FEI: free oestradiol index; HU: Hounsfield unit; LLM: leg lean mass; SHBG: sex hormone-binding globulin; SPPB: short physical performance battery; TT: total testosterone; TT/E2: ratio of total testosterone to oestradiol.

**Table 2.** Participant characteristics from longitudinal analysis.

| Participant Characteristics | n | Baseline | Follow up | Change | p-value |
| --- | --- | --- | --- | --- | --- |
| **General** |  |  |  |  |  |
| Age (years) | 83 | 64.41 ± 10.99 | 69.24 ± 10.95 | 4.83 ± 0.88 | <0.01* |
| **Sex hormone levels** |  |  |  |  |  |
| E2 (pmol/L) | 83 | 50.97 ± 93.99 | 25.1 ± 24.9 | -25.87 ± 87.61 | <0.01* |
| FEI | 83 | 0.08 ± 0.14 | 0.05 ± 0.08 | -0.03 ± 0.14 | 0.08 |
| TT (nmol/L) | 83 | 0.76 ± 0.59 | 0.76 ± 0.43 | 0 ± 0.45 | 0.95 |
| BioT (nmol/L) | 83 | 0.13 ± 0.09 | 0.13 ± 0.09 | 0 ± 0.06 | 0.72 |
| SHBG (nmol/L) | 83 | 72.82 ± 38.55 | 70.09 ± 40.49 | -2.73 ± 33.14 | 0.46 |
| TT/E2 ratio | 83 | 41.09 ± 29.77 | 45.86 ± 30.02 | 4.77 ± 21.83 | 0.05 |
| **Body composition** |  |  |  |  |  |
| BMI (kg/m<sup>2</sup>) | 83 | 27.23 ± 4.62 | 27.34 ± 5.13 | 0.1 ± 1.84 | 0.61 |
| ALM (kg) | 83 | 17.69 ± 2.55 | 17.65 ± 2.53 | -0.04 ± 1.48 | 0.80 |
| Total body fat (%) | 83 | 39.87 ± 6.8 | 39.47 ± 7.98 | -0.4 ± 3.55 | 0.31 |
| Relative appendicular lean mass (kg/kg total fat) | 83 | 0.65 ± 0.22 | 0.69 ± 0.32 | 0.04 ± 0.19 | 0.07 |
| Thigh muscle CSA (cm<sup>2</sup>) | 61 | 91.87 ± 19.07 | 87.67 ± 19.89 | -4.19 ± 6.13 | <0.01* |
| Thigh subcutaneous fat area (cm<sup>2</sup>) | 61 | 113.14 ± 40.96 | 111.27 ± 43.27 | -1.87 ± 19.44 | 0.11 |
| Thigh intramuscular fat area (cm<sup>2</sup>) | 61 | 11.57 ± 4.11 | 12.2 ± 4.87 | 0.54 ± 2.58 | 0.46 |
| Thigh muscle percentage (%) | 61 | 39.57 ± 6.89 | 38.87 ± 7.16 | -0.7 ± 3.36 | 0.11 |
| Thigh muscle density (HU) | 61 | 50.56 ± 3.44 | 48.56 ± 4.28 | -2 ± 2.81 | <0.01* |
| **Muscle function** |  |  |  |  |  |
| Hand grip strength (kg) | 83 | 24.39 ± 6.08 | 24.05 ± 6.76 | -0.34 ± 4.8 | 0.52 |
| Hamstring peak torque (Nm) |  |  |  |  |  |
| 180d/s | 83 | 50.5 ± 15.71 | 44.44 ± 11.86 | -6.06 ± 11.79 | <0.01* |
| 30d/s | 83 | 45.67 ± 14.58 | 44.57 ± 14.16 | -1.11 ± 12.72 | 0.43 |
| Quadriceps peak torque (Nm) |  |  |  |  |  |
| 180d/s | 83 | 62.62 ± 20.49 | 57.58 ± 17.09 | -5.04 ± 17.09 | <0.01* |
| 30d/s | 83 | 103.66 ± 31.29 | 95.12 ± 29.53 | -8.54 ± 19.8 | <0.01* |
| Hamstring relative peak torque to LLM (Nm/kg) |  |  |  |  |  |
| 180d/s | 83 | 3.69 ± 0.92 | 3.28 ± 0.72 | -0.41 ± 0.9 | <0.01* |
| 30d/s | 83 | 3.35 ± 0.93 | 3.28 ± 0.92 | -0.06 ± 0.97 | 0.56 |
| Quadriceps relative peak torque to LLM (Nm/kg) |  |  |  |  |  |
| 180d/s | 83 | 4.58 ± 1.29 | 4.24 ± 1.05 | -0.35 ± 1.42 | 0.03* |
| 30d/s | 83 | 7.58 ± 1.91 | 6.96 ± 1.76 | -0.62 ± 1.67 | 0.01* |
| Hamstring relative peak torque to muscle CSA (Nm/cm<sup>2</sup>) |  |  |  |  |  |
| 180d/s | 61 | 0.56 ± 0.12 | 0.51 ± 0.11 | -0.05 ± 0.12 | <0.01* |
| 30d/s | 61 | 0.5 ± 0.11 | 0.52 ± 0.13 | 0.01 ± 0.13 | 0.51 |
| Quadriceps relative peak torque to muscle CSA (Nm/cm<sup>2</sup>) |  |  |  |  |  |
| 180d/s | 61 | 0.69 ± 0.17 | 0.68 ± 0.16 | -0.01 ± 0.18 | 0.78 |
| 30d/s | 61 | 1.17 ± 0.25 | 1.11 ± 0.26 | -0.05 ± 0.2 | 0.05* |
| **Physical function** |  |  |  |  |  |
| 400m walk time (s) | 50 | 261.19 ± 40.41 | 284.62 ± 63.5 | 23.44 ± 35.62 | <0.01* |
| Time taken to complete 5 chair stands (s) | 74 | 9.83 ± 2.66 | 9.47 ± 3.03 | -0.36 ± 2.29 | 0.18 |
| Mobility disability (%(n)) | 83 |  |  |  |  |
| No (SPPB > 9) |  | 97.59 (81) | 93.98 (78) | - | - |
| Yes (SPPB ≤ 9) |  | 2.41 (2) | 6.02 (5) | - | - |
| Falls in last 12 months (% (n)) | 83 |  |  |  |  |
| No falls |  | 72.29 (60) | 69.88 (58) | - | - |
| Falls |  | 27.71 (23) | 30.12 (25) | - | - |

Physical function measures were only assessed in females ≥ 50 years old. *p*-value >0.05 indicates a statistically significant difference between baseline and follow up. ALM: appendicular lean mass; BioT: bioavailable testosterone; BMI: body mass index; CSA: cross-sectional area; E2: oestradiol; FEI: free oestradiol index; HU: Hounsfield unit; LLM: leg lean mass; SHBG: sex hormone-binding globulin; SPPB: short physical performance battery; TT: total testosterone; TT/E2: ratio of total testosterone to oestradiol. Data presented as mean ± SD. \* Indicates a significant change from baseline to follow-up (*p*-value < 0.05).

### Cross-sectional analysis

Adjusted cross-sectional associations between circulating sex hormones and skeletal muscle outcome measures in females between 24-89 years are displayed in Figure 1-3. Amongst the body composition measures, circulating E2 and FEI were positively associated with relative ALM (kg, *p*<0.05). BioT was positively associated with absolute ALM (kg, *p*<0.05) but negatively associated with relative ALM (kg, *p*<0.05). E2, FEI, and TT were negatively associated with total body fat percentage (*p*<0.05), but there was a positive association for BioT (*p*<0.05). E2 and FEI were positively associated with thigh muscle percentage (*p*<0.05), and TT/E2 ratio was negatively associated with thigh muscle CSA (cm^2^, *p*<0.05). BioT was positively associated with subcutaneous and intramuscular fat (kg, *p*<0.05), while E2 was negatively associated with subcutaneous fat (kg, *p*<0.05). In muscle function models, TT/E2 ratio was negatively associated with hamstring peak torque (Nm) at 180°/s (*p<0*.05) and BioT was positively associated with 400 m walk time (s, *p*<0.05). The binomial linear regression results are displayed in Table S2. There were no significant associations between any hormones and indices of mobility disability or falls in females over 50 years (n=238).

**Fig 1.**
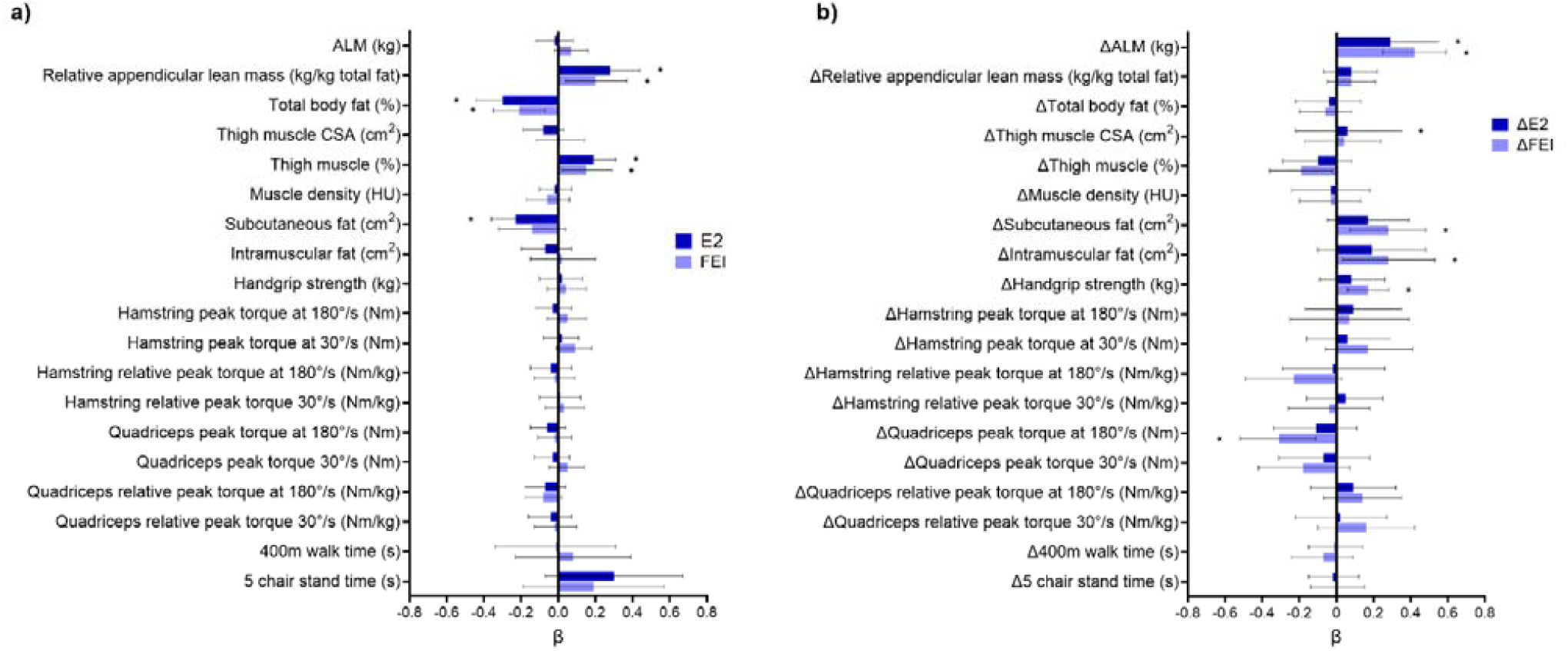
A) Adjusted cross-sectional associations between serum oestradiol (E2) and free oestrogen index (FEI) and muscle outcomes. B) Adjusted longitudinal associations between the change in serum E2 and FEI and the change in muscle outcomes across 4-6 years in females ≥ 50 years old. Multiple regression models were adjusted for age, ethnicity, height, ACCI, and total physical activity levels in the past two weeks. Physical function measures were only assessed in females ≥ 50 years old. Relative hamstring and quadriceps peak torque (Nm/kg) have been normalised to leg lean mass (kg). ALM: appendicular lean mass; CSA: cross-sectional area; HU: Hounsfield unit. Data presented as beta coefficient (β) and 95% confidence interval. * Indicates a significant association (*p*-value < 0.05)

**Fig 2.**
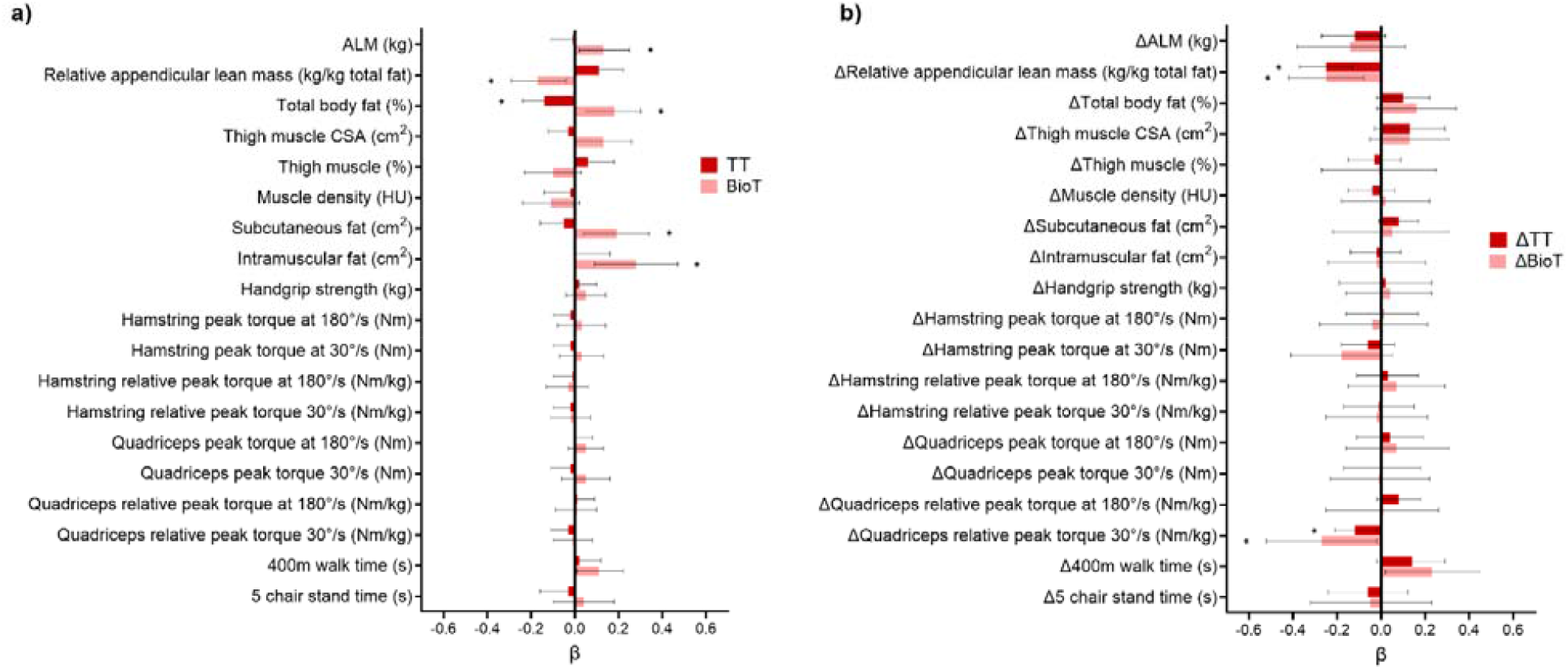
A) Adjusted cross-sectional associations between total serum testosterone (TT) and bioavailable testosterone (BioT) and muscle outcomes. B) Adjusted longitudinal associations between the change in TT and BioT and the change in muscle outcomes across 4-6 years in females ≥ 50 years old. Multiple regression models were adjusted for age, ethnicity, height, ACCI, and total physical activity levels in the past two weeks. Physical function measures were only assessed in females ≥ 50 years old. Relative hamstring and quadriceps peak torque (Nm/kg) have been normalised to leg lean mass (kg). ALM: appendicular lean mass; CSA: cross-sectional area; HU: Hounsfield unit. Data presented as beta coefficient (β) and 95% confidence interval. * Indicates a significant association (*p*-value < 0.05)

**Fig 3.**
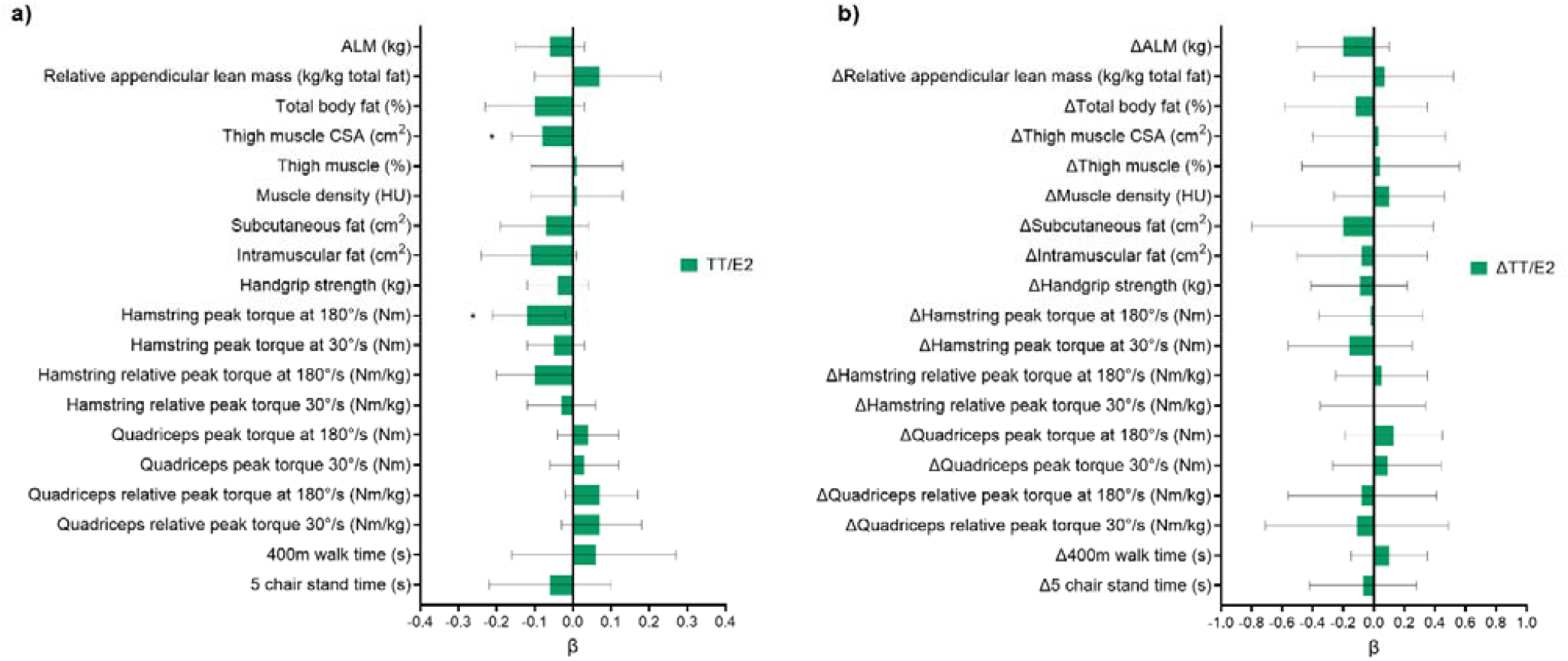
A) Adjusted cross-sectional associations between the total testosterone to total oestradiol (TT/E2) ratio and muscle outcomes. B) Adjusted longitudinal associations between the change in serum TT/E2 ratio and the change in muscle outcomes across 4-6 years in females ≥ 50 years old. Multiple regression models were adjusted for age, ethnicity, height, ACCI, and total physical activity levels in the past two weeks. Physical function measures were only assessed in females ≥ 50 years old. Relative hamstring and quadriceps peak torque (Nm/kg) have been normalised to leg lean mass (kg). ALM: appendicular lean mass; CSA: cross-sectional area; HU: Hounsfield unit. Data presented as beta coefficient (β) and 95% confidence interval. * Indicates a significant association (*p*-value < 0.05)

Cross-sectional associations between menopausal status, hormonal supplementation and muscle outcomes are displayed in Table S3. Postmenopausal status was positively associated with total body fat (%, *p*<0.05), subcutaneous fat (kg, *p*<0.05), and intramuscular fat (kg, *p*<0.05), and negatively associated with relative ALM (kg, *p*<0.05) and thigh muscle CSA (cm^2^, *p*<0.05). Amongst postmenopausal females, use of HRT was positively associated with handgrip strength (kg, *p*<0.05). In premenopausal females, use of the HCP was positively associated with relative quadriceps strength normalised to leg lean mass at 30°/s (Nm/kg, *p*<0.05). It was also negatively associated with muscle density (HU, *p*<0.05), absolute and relative hamstring strength (Nm and Nm/kg, *p*<0.05), and relative quadriceps strength normalised to thigh muscle CSA at 30°/s (Nm/cm^2^, *p*<0.05).

Where E2/FEI and BioT were found to have opposite associations with a muscle outcome, both hormones were included in the model. BioT was added to the cross-sectional models investigating the relationship between E2 and relative ALM, total body fat percentage, and subcutaneous fat, and the relationship between FEI and relative ALM and total body fat. In all models, oestradiol (E2 or FEI) remained significantly associated positively with relative ALM and negatively with body fat percentage and subcutaneous fat, while BioT remained significantly associated in the opposite direction (*p<0*.05; Table S4).

### Longitudinal analysis

Adjusted longitudinal associations between the change in circulating sex hormones and the change in skeletal muscle outcome measures in females over 50 years are displayed in Figures 1-3. Regarding body composition, ΔE2 and ΔFEI were positively associated with the change in ALM (kg, *p*<0.05). ΔTT and ΔBioT were negatively associated with the change in relative ALM (kg, *p*<0.05). ΔFEI was negatively associated with the change in thigh muscle percentage (*p*<0.05), and positively associated with the change in subcutaneous and intramuscular fat (kg, *p*<0.05). Regarding muscle function, ΔFEI was positively associated with the change in handgrip strength (kg, *p*<0.05), but negatively associated with the change in quadriceps strength (Nm) at 180°/s (*p*<0.05). Similarly, ΔTT and ΔBioT were negatively associated with the change in relative quadriceps strength at 30°/s when normalised to leg lean mass (Nm/kg, *p*<0.05). Finally, ΔBioT was positively associated with the change in 400 m walk time (s, *p*<0.05).

Longitudinal associations between baseline menopausal status, HRT use and the change in muscle outcomes across 4-6 years in females over 50 years old are displayed in Table S5. Postmenopausal status at baseline was positively associated with the change in ALM (kg, *p*<0.05) and thigh muscle CSA (cm^2^, *p*<0.05). Being a current HRT user at baseline was negatively associated with the change in handgrip strength (kg, *p*<0.05), while previous use of HRT was positively associated with the change in intramuscular fat (kg, *p*<0.05) and relative hamstring strength normalised to muscle thigh CSA (Nm/cm^2^, *p*<0.05), and negatively associated with the change in thigh muscle CSA (cm^2^, *p*<0.05).

## Discussion

To the best of the authors knowledge, this is the first study to 1) map cross-sectional associations between sex hormones and muscle outcomes in a wide continuum of ages across the adult lifespan, and 2) investigate how changes to circulating sex hormones beyond 50 years of age associate with changes to muscle mass and function over 4-6 years.

### Sex hormones and body composition

#### Oestradiol

This study highlighted a positive cross-sectional association between serum E2 levels (total and FEI) and measures of muscle mass (relative ALM and thigh muscle percentage) in females across the lifespan, independent of age. Additionally, both total serum E2 and FEI were negatively associated with total body fat percentage and thigh subcutaneous fat, suggesting a potential role for E2 in maintaining the proportion of muscle mass throughout ageing. This is line with previous mechanistic work in animals highlighting an anabolic role of E2 in skeletal muscle [12]. Interestingly, this contradicts with many previous cross-sectional studies investigating samples of postmenopausal females, which demonstrate no associations between serum oestradiol or FEI and muscle mass [13]. This is likely due to the inclusion of postmenopausal females that, by definition, have very low serum E2 concentrations, so there is unlikely to be large variation between participants. In contrast, the current cohort spans a wide continuum of adulthood (24 to 89 years) and E2 concentrations (5 to 1872 pmol/L), thereby allowing us to understand how large age-related fluctuations in oestrogens impact the regulation of muscle mass over the lifespan.

In the longitudinal analysis of females over 50 years old, the change in serum E2 and FEI across the 5-year follow up was positively associated with the change in absolute ALM, suggesting females with a larger decline in serum E2 and FEI have a greater decline in muscle mass. This further emphasises a potential role for total and free E2 in anabolic regulation, in line with the cross-sectional findings. The only longitudinal study to have previously investigated this found no association between serum E2 and the loss of ALM over 3 years in 49 postmenopausal females [14]. However, baseline hormone concentrations were used in the model, as opposed to the change in hormone concentration from baseline to follow up. This suggests that it may not be baseline E2 concentration, but the magnitude of change in E2 concentration from baseline, that determines the extent of muscle loss post-menopause. It is therefore important to understand the temporal nature of the relationship between E2 and skeletal muscle, where the population is displaying rapid changes in both hormone concentrations and muscle mass and function.

Surprisingly, the change in serum FEI was negatively associated with the change in thigh muscle percentage, and positively associated with the change in thigh subcutaneous and intramuscular fat. Together this suggests that a decline in serum FEI coincides with an increased relative proportion of muscle in the thigh, potentially due to declines in both subcutaneous and intramuscular fat content, rather than an inverse relationship between FEI and muscle mass. We suggest this is the case, as the change in FEI was positively associated with absolute ALM. Although, DXA measures of ALM do not appropriately distinguish muscle from intramuscular fat, which may have exaggerated this positive association, highlighting the importance of including direct measures of muscle and fat infiltration from CT scans.

#### Testosterone

In the cross-sectional analysis, serum TT concentrations were negatively associated with total body fat percentage and no other compositional measures. This in line with previous studies showing a lack of association between serum TT and lean mass or muscle CSA in both pre-[11, 19] and postmenopausal [20-23] females, although this is the first study to investigate the association across the entire adult lifespan. Instead, bioavailable (non-SHBG bound) or free (non-protein bound) forms of testosterone appear to be more relevant in this context, as many studies have reported a positive association with lean mass [11, 20, 21, 23-26] or muscle CSA [27-29].

In line with these studies, BioT concentration was positively associated with absolute ALM. However, it was also positively associated with measures of fat mass (total body fat percentage, and thigh subcutaneous and intramuscular fat), explaining the negative association between BioT and relative ALM; an increase in fat tissue corresponds to a reduction in relative ALM, as relative ALM is indexed to total body fat mass. While change in muscle and fat mass across 4-6 years of ageing may occur in the same direction (i.e. both gained or lost), the resulting relative proportion of each tissue differs. A similar trend was noted in the longitudinal analysis, as both the change in TT and BioT from baseline to follow up were negatively associated with the change in relative ALM.

### Sex hormones and muscle function

#### Oestradiol

No significant cross-sectional associations were present between serum E2 or FEI and any measure of muscle function. This contrasts with Pöllanen et al. [30] who found a positive association between serum E2 and relative quadriceps femoris strength in an albeit much smaller sample (n=25) of pre- and postmenopausal females. Importantly, age was not included as a confounding variable, therefore these findings may just reflect the loss of both serum E2 and muscle strength with age. Indeed, when age was included as a covariate, Bochud et al. [31] found no cross-sectional association between urinary E2 concentrations and handgrip strength in a sample of 366 females between 18 and 90 years of age.

The change in serum FEI was significantly and positively associated with the change in handgrip strength across 4-6 years, suggesting that a greater decline in free oestradiol is related to a greater decline in handgrip strength, a common proxy for overall muscle and physical function [32]. In contrast, the change in serum FEI was negatively associated with the change in absolute quadriceps strength. This finding is unexpected but may indicate the presence of other confounding factors that are not yet identified. Rolland et al. [14] found no association between serum E2 and the loss of muscle strength over 3 years in postmenopausal females, but higher serum oestrone (E1) was similarly associated with a greater loss of muscle strength. E1 is an endogenous form of oestrogen which becomes predominant after the menopause-related decline of serum E2 [30]. While less potent than E2, its role within the muscle remains largely understudied, highlighting a target for future research, particularly in postmenopausal females.

#### Testosterone

Similarly to E2, both serum TT and BioT were not cross-sectionally associated with any measures of muscle strength. This is in line with other studies that investigated younger [11, 19, 27], older [33] or both pre- and postmenopausal [30] females. To our knowledge, this is the first study to investigate longitudinal associations between serum testosterone and muscle function in ageing females. The change in both TT and BioT levels were negatively associated with the change in relative quadriceps strength, suggesting that an increase in serum testosterone concentration coincides with a decrease in lower limb strength. While this is unexpected, it is further supported by the positive cross-sectional and longitudinal associations between BioT and gait speed. This suggests that greater concentrations of, or changes to, circulating bioavailable testosterone associate with worse physical function in the lower limbs of ageing females. This is surprising as testosterone supplementation in postmenopausal females can elicit anabolic effects in skeletal muscle [34], although basal molecular regulation of muscle by testosterone has not yet been investigated in females.

### The TT/E2 ratio and skeletal muscle ageing

When investigating their effects within skeletal muscle, circulating sex hormones have most often been considered individually, but their production is closely linked due to common biosynthetic pathways. For example, testosterone is converted to oestradiol via the enzyme aromatase; a process that can be quantitatively assessed by the TT/E2 ratio, where a greater TT/E2 ratio indicates less synthesis of E2 from testosterone, and therefore lower aromatase activity [35]. In premenopausal females, the ovaries are responsible for ∼95% of circulating oestradiol production, but the peripheral conversion of testosterone to E2 becomes more physiologically relevant after menopause when the ovaries cease E2 production [35, 36]. We observed a negative cross-sectional association between the TT/E2 ratio and muscle mass (thigh muscle CSA) and strength (hamstring peak torque), suggesting that a lower conversion rate of testosterone to E2 is concomitant to poorer muscle quality across ageing. Further research should be conducted to understand the contribution of varying oestrogen synthesis pathways in the regulation of skeletal muscle throughout ageing in females.

### Limitations

While this study provides novel insights into the relationship between circulating sex hormones and skeletal muscle ageing, it is not without limitations. Menstrual cycle phase has not been controlled when scheduling premenopausal participant visits. Common measures of acute skeletal muscle function are unlikely to be altered according to menstrual cycle phase [37], however, circulating E2 levels can vary from 70-1500 pmol/L between the early follicular and ovulatory phase [38]. Therefore, depending on the phase in which the blood sample was collected, the relationship between circulating E2 and skeletal muscle outcomes may significantly differ. Future studies should test all premenopausal participants within the early follicular phase as previously described [39], where serum E2 is at its lowest or ‘basal’ level.

Secondly, premenopausal participants provided information on HCP use, but many other forms of hormonal contraception are commonly used and provide external doses of oestrogen or progesterone, including the intrauterine devices, implant, injections and skin patches. Therefore, a more comprehensive understanding of exogenous sex hormone profiles is required to improve reliability of the findings.

Finally, approximately one third of participants in this study were non-white, suggesting that these findings are representative of a range of ethnicities. However, due to the inclusion criteria of the BLSA, participants are likely healthier than the general population. Therefore, the generalisability of these findings to females with multiple chronic health conditions is questionable.

## Conclusion

This study has demonstrated novel and divergent cross-sectional and longitudinal associations between circulating sex hormones and skeletal muscle outcomes in females across the ageing continuum. This emphasises the need to consider the temporal aspect of sex hormone regulation in ageing female muscle, particularly across the transitional period from a high to low state of circulating oestrogen. Future research should aim to identify molecular mechanisms of oestrogen action in human female skeletal muscle, to determine how ageing-induced oestrogen deficiency contributes to a loss of muscle mass and function in the later decades of life.

## Data Availability

All data produced in the present work are contained in the manuscript.

https://www.blsa.nih.gov/blsa-data-use

## Additional Information

### Ethical Standards

Written informed consent was provided by all participants at every visit and the BSLA protocol was approved by the National Institute of Health (NCT00233272). This study (project number: 2023-029) has been declared exempt from ethical review on 07/02/2023 by the Deakin University Human Research Ethics Committee (DUHREC) as it comprises only of secondary data analysis.

