## Supplementary File for "Associations Between Female Sex Hormones and Skeletal Muscle Ageing: The Baltimore Longitudinal Study of Aging"

**Table S1.** Menopausal status and hormonal supplementation of females in the cross-sectional and longitudinal analysis.

| **Hormonal characteristic** | **Cross-sectional** | | **Longitudinal (Baseline)** | | **Longitudinal (Follow-up)** | |
| --- | --- | --- | --- | --- | --- | --- |
|  | *n* | *% (n)* | *n* | *% (n)* | *n* | *% (n)* |
| **Menopausal Status** | 295 |  | 81 |  | 82 |  |
| *Premenopausal* |  | 29.83 (88) |  | 14.81 (12) |  | 6.10 (5) |
| *Postmenopausal* |  | 64.07 (189) |  | 76.54 (62) |  | 86.59 (71) |
| *Unsure* |  | 6.10 (18) |  | 8.64 (7) |  | 7.32 (6) |
| **Oestrogen HRT use amongst postmenopausal females:** | 186 |  | 60 |  | 67 |  |
| *Currently* |  | 16.58 (31) |  | 12.50 (10) |  | 5.97 (4) |
| *Previously* |  | 39.57 (74) |  | 46.67 (28) |  | 47.76 (32) |
| *Never* |  | 43.32 (81) |  | 38.33 (23) |  | 46.27 (31) |
| **HCP use in premenopausal females:** | 85 |  | 12 |  | 5 |  |
| *Currently* |  | 12.94 (11) |  | 0.00 (0) |  | 0.00 (0) |
| *Previously* |  | 68.24 (58) |  | 83.33 (10) |  | 80.00 (4) |
| *Never* |  | 18.82 (16) |  | 16.67 (2) |  | 20.00 (1) |

All available data is presented (n values vary as these measures were not always collected in the BLSA). HCP: hormonal contraceptive pill; HRT: hormonal replacement therapy.

**Table S2.** Cross-sectional associations between serum sex hormones and dichotomous physical function outcomes in females ≥ 50 years old.

| **Outcome** | **n** | **Hormone** | ***Odds ratio*** | ***95% CI*** | ***P*** |
| --- | --- | --- | --- | --- | --- |
| Faller | 238 | E2 | 0.90 | 0.72 - 1.13 | 0.36 |
|  |  | FEI | 0.81 | 0.52 - 1.24 | 0.33 |
|  |  | TT | 1.01 | 0.99 - 1.03 | 0.33 |
|  |  | BioT | 1.02 | 0.91 - 1.14 | 0.71 |
|  |  | SHBG | 1.00 | 0.99 - 1.01 | 0.72 |
|  |  | TT/E2 ratio | 1.01 | 1.00 - 1.02 | 0.27 |
| Mobility disability | 238 | E2 | 0.47 | 0.05 - 4.25 | 0.51 |
|  |  | FEI | 0.87 | 0.31 - 2.46 | 0.79 |
|  |  | TT | 0.98 | 0.87 - 1.10 | 0.72 |
|  |  | BioT | 1.17 | 0.94 - 1.47 | 0.17 |
|  |  | SHBG | 0.91 | 0.83 - 1.01 | 0.07 |
|  |  | TT/E2 ratio | 1.00 | 0.95 - 1.05 | 0.93 |

BioT: bioavailable testosterone; E2: oestradiol; FEI: free oestradiol index; SHBG: sex hormone-binding

globulin; TT: total testosterone; TT/E2: ratio of total testosterone to oestradiol. * Indicates statistical

significance (*p*<0.05).

**Table S3.** Adjusted cross-sectional associations between menopausal and exogenous hormonal supplementation status and muscle outcomes.

| **Outcome variable** |  | **Postmenopausal Status** | | | | **HRT use (postmenopausal females only)** | | | | **HCP use (premenopausal females only)** | | | |
| --- | --- | --- | --- | --- | --- | --- | --- | --- | --- | --- | --- | --- | --- |
|  |  | **n** | **B** | ***R^2^*** | ***P*** | **n** | **B** | ***R^2^*** | ***P*** | **n** | **B** | ***R^2^*** | ***P*** |
| ***Body composition*** | ALM (kg) | 295 | -0.18 | 0.31 | 0.61 | 187 | 0.20 | 0.29 | 0.68 | 86 | 0.68 | 0.33 | 0.25 |
|  | Relative appendicular lean mass (kg/kg total fat) | 295 | -0.12 | 0.09 | **<0.01*** | 187 | 0.02 | 0.08 | 0.71 | 86 | -0.21 | 0.23 | 0.11 |
|  | Total body fat (%) | 295 | 3.42 | 0.09 | **<0.01*** | 187 | -1.01 | 0.10 | 0.47 | 86 | 5.35 | 0.21 | 0.11 |
|  | Thigh muscle CSA (cm^2^) | 219 | -1.17 | 0.44 | 0.66 | 144 | 1.85 | 0.46 | 0.56 | 55 | 0.62 | 0.28 | 0.94 |
|  | Thigh muscle (%) | 219 | -4.76 | 0.24 | **<0.01*** | 144 | -0.23 | 0.09 | 0.90 | 55 | -8.42 | 0.29 | 0.12 |
|  | Muscle density (HU) | 219 | -0.97 | 0.26 | 0.11 | 144 | 0.30 | 0.15 | 0.70 | 55 | -1.96 | 0.12 | **0.02*** |
|  | Subcutaneous fat (cm^2^) | 219 | 25.56 | 0.13 | **<0.01*** | 144 | 2.01 | 0.14 | 0.85 | 55 | 50.08 | 0.25 | 0.08 |
|  | Intramuscular fat (cm^2^) | 219 | 1.66 | 0.06 | **0.02*** | 144 | 1.25 | 0.06 | 0.28 | 55 | -0.60 | 0.18 | 0.73 |
| ***Muscle Function*** | Handgrip strength (kg) | 295 | -0.84 | 0.42 | 0.32 | 187 | 2.03 | 0.42 | **0.01*** | 86 | -2.00 | 0.20 | 0.27 |
|  | Hamstring peak torque (Nm) |  |  |  |  |  |  |  |  |  |  |  |  |
|  | *180d/s* | 295 | 2.56 | 0.33 | 0.14 | 187 | 0.95 | 0.33 | 0.71 | 86 | -7.32 | 0.22 | **<0.05*** |
|  | *30d/s* | 295 | 0.85 | 0.35 | 0.62 | 187 | 0.95 | 0.33 | 0.68 | 86 | -8.52 | 0.20 | 0.06 |
|  | Hamstring relative peak torque (Nm/kg LLM) |  |  |  |  |  |  |  |  |  |  |  |  |
|  | *180d/s* | 295 | 0.25 | 0.14 | 0.06 | 187 | -0.03 | 0.18 | 0.86 | 86 | -0.67 | 0.13 | **0.02*** |
|  | *30d/s* | 295 | 0.14 | 0.18 | 0.25 | 187 | -0.03 | 0.17 | 0.83 | 86 | -0.69 | 0.09 | **0.02*** |
|  | Hamstring relative peak torque (Nm/cm^2^) |  |  |  |  |  |  |  |  |  |  |  |  |
|  | *180d/s* | 219 | 0.03 | 0.12 | 0.18 | 144 | -0.01 | 0.12 | 0.77 | 55 | -0.06 | 0.27 | 0.14 |
|  | *30d/s* | 219 | 0.01 | 0.11 | 0.67 | 144 | -0.03 | 0.08 | 0.25 | 55 | -0.06 | 0.23 | 0.24 |
|  | Quadriceps peak torque (Nm) |  |  |  |  |  |  |  |  |  |  |  |  |
|  | *180d/s* | 295 | 2.14 | 0.43 | 0.41 | 187 | -0.42 | 0.46 | 0.89 | 86 | -10.65 | 0.25 | 0.16 |
|  | *30d/s* | 295 | 1.65 | 0.36 | 0.66 | 187 | 1.92 | 0.36 | 0.74 | 86 | -17.50 | 0.22 | 0.06 |
|  | Quadriceps relative peak torque (Nm/kg LLM) |  |  |  |  |  |  |  |  |  |  |  |  |
|  | *180d/s* | 295 | 0.03 | 0.25 | 0.32 | 187 | -0.17 | 0.36 | 0.43 | 86 | -0.94 | 0.18 | 0.08 |
|  | *30d/s* | 295 | 0.06 | 0.18 | 0.11 | 187 | -0.13 | 0.26 | 0.71 | 86 | 1.49 | 0.13 | **0.02*** |
|  | Quadriceps relative peak torque (Nm/cm^2^) |  |  |  |  |  |  |  |  |  |  |  |  |
|  | *180d/s* | 219 | 0.24 | 0.30 | 0.21 | 144 | -0.02 | 0.26 | 0.51 | 55 | -0.13 | 0.38 | 0.05 |
|  | *30d/s* | 219 | 0.25 | 0.22 | 0.33 | 144 | -0.04 | 0.16 | 0.46 | 55 | -0.24 | 0.31 | **0.03*** |
| ***Physical Function*** | 400m walk time (s) | 146 | 7.80 | 0.30 | 0.45 | 122 | 6.44 | 0.27 | 0.49 | - | - | - | - |
|  | 5 chair stand time (s) | 186 | 0.46 | 0.09 | 0.33 | 152 | 0.15 | 0.08 | 0.81 | - | - | - | - |

Multiple regression models were adjusted for age, ethnicity, height, ACCI, and total physical activity levels in the past two weeks. Physical function measures were only assessed in females ≥ 50 years old. ALM: appendicular lean mass; CSA: cross-sectional area; HCP: hormonal contraceptive pill; HRT: hormonal replacement therapy; HU: Hounsfield unit; LLM: leg lean mass. * Indicates statistical significance (*p*<0.05).

**Table S4:** Multiple hormone cross-sectional linear regression models.

| **Outcome Variable** | **n** | **Hormone** | ***R^2^*** | **β** | ***P*** |
| --- | --- | --- | --- | --- | --- |
| Relative ALM (kg/kg total body fat) | 319 | E2 | 0.19 | 0.26 | **<0.01*** |
|  |  | BioT |  | -0.15 | **0.02*** |
|  |  | FEI | 0.17 | 0.22 | **<0.01*** |
|  |  | BioT |  | -0.18 | **<0.01*** |
| Body fat percentage (%) | 319 | E2 | 0.17 | -0.29 | **<0.01*** |
|  |  | BioT |  | 0.16 | **0.01*** |
|  |  | FEI | 0.15 | -0.23 | **<0.01*** |
|  |  | BioT |  | 0.19 | **<0.01*** |
| Subcutaneous fat (kg) | 239 | E2 | 0.16 | -0.22 | **<0.01*** |
|  |  | BioT |  | 0.17 | **<0.03*** |

Where significant and opposing associations were detected for serum oestradiol (E2 or FEI) and testosterone (TT or

BioT), both hormones were added to the model to determine if they remained significantly and independently associated

with the outcome variable. ALM: appendicular lean mass; BioT: bioavailable testosterone; E2: oestradiol; FEI: free

oestradiol index. * Indicates statistical significance (*p*<0.05).

**Table S5.** Adjusted longitudinal associations between baseline menopausal and exogenous hormonal supplementation status and the change in muscle outcomes across 4-6 years in females ≥ 50 years old.

| **Outcome variable** |  | **Postmenopausal status** | | | | **HRT user at baseline** | | | | **Previous HRT user at baseline** | | | |
| --- | --- | --- | --- | --- | --- | --- | --- | --- | --- | --- | --- | --- | --- |
|  |  | **n** | **B** | ***R^2^*** | ***P*** | **n** | **B** | ***R^2^*** | ***P*** | **n** | **B** | ***R^2^*** | ***P*** |
| ***Body composition*** | ΔALM (kg) | 81 | 1.19 | 0.20 | **<0.01*** | 61 | 0.08 | 0.13 | 0.86 | 61 | -0.20 | 0.14 | 0.64 |
|  | ΔRelative appendicular lean mass (kg/kg total fat) | 81 | 0.12 | 0.15 | **0.01*** | 61 | 0.06 | 0.17 | 0.58 | 61 | -0.08 | 0.20 | 0.13 |
|  | ΔTotal body fat (%) | 81 | -1.16 | 0.09 | 0.24 | 61 | 1.67 | 0.17 | 0.27 | 61 | 0.66 | 0.16 | 0.52 |
|  | ΔThigh muscle CSA (cm^2^) | 59 | 3.18 | 0.18 | **<0.05*** | 41 | -0.70 | 0.23 | 0.72 | 41 | -4.01 | 0.32 | **0.04*** |
|  | ΔThigh muscle (%) | 59 | 0.40 | 0.05 | 0.65 | 41 | 0.96 | 0.13 | 0.59 | 41 | -1.95 | 0.18 | 0.10 |
|  | ΔMuscle density (HU) | 59 | 0.22 | 0.07 | 0.75 | 41 | -1.48 | 0.10 | 0.10 | 41 | -0.42 | 0.07 | 0.67 |
|  | ΔSubcutaneous fat (cm^2^) | 59 | 5.84 | 0.14 | 0.43 | 41 | -3.15 | 0.28 | 0.68 | 41 | 6.07 | 0.29 | 0.41 |
|  | ΔIntramuscular fat (cm^2^) | 59 | 0.34 | 0.06 | 2.65 | 41 | -1.89 | 0.11 | 0.20 | 41 | 1.93 | 0.17 | **0.04*** |
| ***Muscle Function*** | ΔHandgrip strength (kg) | 81 | 2.72 | 0.07 | 0.06 | 61 | -3.96 | 0.12 | **0.04*** | 61 | 1.88 | 0.08 | 0.15 |
|  | ΔHamstring peak torque (Nm) |  |  |  |  |  |  |  |  |  |  |  |  |
|  | *180d/s* | 81 | 0.51 | 0.06 | 0.88 | 61 | -2.47 | 0.04 | 0.44 | 61 | 5.51 | 0.08 | 0.08 |
|  | *30d/s* | 81 | 1.80 | 0.04 | 0.65 | 61 | -2.46 | 0.05 | 0.57 | 61 | 3.30 | 0.07 | 0.32 |
|  | ΔHamstring relative peak torque (Nm/kg LLM) |  |  |  |  |  |  |  |  |  |  |  |  |
|  | *180d/s* | 81 | -0.25 | 0.11 | 0.30 | 61 | -0.16 | 0.07 | 0.56 | 61 | 0.42 | 0.12 | 0.09 |
|  | *30d/s* | 81 | -0.13 | 0.04 | 0.65 | 61 | -0.24 | 0.05 | 0.52 | 61 | 0.25 | 0.06 | 0.35 |
|  | ΔHamstring relative peak torque (Nm/cm^2^) |  |  |  |  |  |  |  |  |  |  |  |  |
|  | *180d/s* | 59 | -0.04 | 0.07 | 0.28 | 41 | -0.01 | 0.03 | 0.75 | 41 | 0.06 | 0.10 | 0.13 |
|  | *30d/s* | 59 | -0.03 | 0.07 | 0.55 | 41 | -0.01 | 0.06 | 0.78 | 41 | 0.09 | 0.20 | **0.02*** |
|  | ΔQuadriceps peak torque (Nm) |  |  |  |  |  |  |  |  |  |  |  |  |
|  | *180d/s* | 81 | -2.82 | 0.12 | 0.54 | 61 | -8.31 | 0.14 | 0.17 | 61 | 4.70 | 0.13 | 0.33 |
|  | *30d/s* | 81 | 3.49 | 0.04 | 0.49 | 61 | 5.30 | 0.02 | 0.55 | 61 | 3.41 | 0.02 | 0.57 |
|  | ΔQuadriceps relative peak torque (Nm/kg LLM) |  |  |  |  |  |  |  |  |  |  |  |  |
|  | *180d/s* | 81 | -0.52 | 0.16 | 0.14 | 61 | -0.56 | 0.17 | 0.25 | 61 | 0.35 | 0.17 | 0.40 |
|  | *30d/s* | 81 | -0.24 | 0.05 | 0.58 | 61 | 0.36 | 0.04 | 0.61 | 61 | 0.30 | 0.04 | 0.56 |
|  | ΔQuadriceps relative peak torque (Nm/cm^2^) |  |  |  |  |  |  |  |  |  |  |  |  |
|  | *180d/s* | 59 | -0.04 | 0.16 | 0.40 | 41 | -0.08 | 0.16 | 0.26 | 41 | 0.06 | 0.16 | 0.36 |
|  | *30d/s* | 59 | -0.07 | 0.08 | 0.24 | 41 | 0.01 | 0.05 | 0.96 | 41 | 0.13 | 0.14 | 0.09 |
| ***Physical Function*** | Δ400m walk time (s) | 48 | -16.47 | 0.51 | 0.03 | 34 | 0.58 | 0.45 | 0.94 | 34 | -15.01 | 0.52 | 0.12 |
|  | Δ5 chair stand time (s) | 72 | 0.40 | 0.07 | 0.59 | 53 | -1.00 | 0.09 | 0.12 | 53 | -0.08 | 0.07 | 0.89 |

Multiple regression models were adjusted for age, ethnicity, height, ACCI, and total physical activity levels in the past two weeks. ALM: appendicular lean mass; CSA: cross-sectional area; HRT: hormonal replacement therapy; HU: Hounsfield unit; LLM: leg lean mass. * Indicates statistical significance (*p*<0.05).
